# Towards the next phase: evaluation of serological assays for diagnostics and exposure assessment

**DOI:** 10.1101/2020.04.23.20077156

**Authors:** Corine H. GeurtsvanKessel, Nisreen M.A. Okba, Zsofia Igloi, Carmen W.E. Embregts, Brigitta M. Laksono, Lonneke Leijten, Janette Rahamat-Langendoen, Johannes P.C. van den Akker, Jeroen J.A. van Kampen, Annemiek A. van der Eijk, Rob S. van Binnendijk, Bart Haagmans, Marion Koopmans

## Abstract

The world is entering a new era of the COVID-19 pandemic in which there is an increasing call for reliable antibody testing. To support decision making on the deployment of serology for either population screening or diagnostics, we present a comprehensive comparison of serological COVID-19 assays. We show that the assay detecting total immunoglobulins against the receptor binding domain of SARS CoV-2, had optimal characteristics for antibody detection in different stages of disease.

## Introduction

The novel severe acute respiratory syndrome coronavirus 2 (SARS-CoV-2) which has emerged in China in late 2019 causes coronavirus disease (COVID-19). The rapid global spread and exponential growth of the first pandemic wave in China, Europe and the US have stretched the limits of the available healthcare and ICU capacity, reaching critical levels in the province where the outbreak originated, and elsewhere. Since the initial notification of an outbreak on December 31st, the global response has transitioned from the initial policy of active case finding and containment to an increasingly complex package of confinement measures including closures of schools, restaurants, shops, implementation of travel restrictions, and physical distancing measures. At present, given the global circulation of SARS-CoV-2, the consensus is that elimination of the virus is no longer feasible, and that longer-term strategies are needed that strike a balance between the economically and socially damaging (near) lockdown approaches and full release of any control measures. There is wide agreement that, in the latter situation, rapid resurgence would be very likely, with modelled epidemic peaks potentially exceeding the current healthcare capacity^1^.

The “exit” strategy is defined as the transition from the current approach, which focuses entirely on flattening the peak of the COVID-19 emergence curve, to the transition phase in which restrictions are gradually lifted. The gradual lifting of control measures will require active surveillance to allow early detection of new cases or clusters, coupled with contact tracing and quarantine, most likely combined with continued physical distancing recommendations and enhanced protection of those at risk from most severe disease. A key knowledge gap is the level and duration of immunity in the population at large and in specific groups, including persons with different clinical severity ^1^. To assess the extent of virus circulation in the community, and the likelihood of protection against a re-infection, there is a crucial need to add serology to the testing algorithms. The required performance of a serological assay will depend on the specific aim of testing which may be either population screening (in the general population or at-risk populations) or diagnostic support. When selecting an appropriate assay for a specific purpose, decision making should include the available knowledge on antibody specificities, kinetics, and functions. We recently showed that antibodies directed against the S1 subunit of the SARS-CoV-2 spike protein and specifically to the receptor binding domain (RBD) within the S1 subunit strongly correlate with virus neutralization^2^. The likelihood of predicting protective antibody responses will thus increase when using either S1 antigens or RBD in the assay. Here, we present a comprehensive comparison of a selection of COVID-19 assays for antibody measurements to support decisions on deployment of serology for population testing and diagnostics. The list of commercial assays offered is growing exponentially ([FINDDX inventory](https://www.finddx.org/covid-19/pipeline/), currently 254 immunoassays are listed). We chose to include commercial assays that 1) preferably target the spike of SARS-CoV-2 2) had received at least European Conformity (CE) marking or other authorization, and 3) ensured sufficient production capacity.

## Methods

### Blood Samples

We used a well-defined specificity panel of 147 serum and plasma samples from persons exposed to a human coronaviruses (HCoV-229E, NL63 or OC43), SARS, MERS, or with a range of other respiratory viruses (*Table 1*) to determine specificity of the assays. Specimen from patients with recent CMV, EBV or *M. pneumoniae* infection were included as these have a high likelihood of causing cross reactivity. To determine the sensitivity, 93 sera from 24 confirmed COVID-19 patients with different levels of disease severity were analyzed. All specimen were stored at −20°C until use.

### Ethical clearance

The use of specimen from the Netherlands was approved by the local medical ethical committee (MEC approval: 2014–414). Serum samples from SARS patients were kindly provided by professor Malik Peiris, Hong Kong University.

### ELISA

ELISAs were selected based on predefined selection criteria like the coating antigen, authorizations for use by various national regulatory agencies, availability of large scale production and available validation data: 1) Wantai SARS-CoV-2 total Ig and IgM ELISA from Beijing Wantai Biological Pharmacy Enterprise Co., Ltd and 2) Anti-SARS-CoV-2 IgG and IgA ELISA assay (EUROIMMUN Medizinische Labordiagnostika AG. ELISAs were performed according to manufacturer’s protocol.

### DiaSorin Liaison XL

The Liaison XL is a semi-automated system using chemiluminescent immunoassay (CLIA) technology for detection of Ab in human samples. The assays were performedfollowing manufacturer’s protocol.

### Rapid antibody test

The rapid tests we evaluated are 1) Rapid SARS -CoV-2 Antibody (IgM/IgG) Test from InTec utilizing the nucleocapsid protein as antigen (Test of lot S2020021505), 2) the qSARS-CoV-2 IgG/IgM Cassette Rapid Test (GICA) from Cellex Inc. utilizing both the spike and the nucleocapsid protein (Test lot 0200311WI5513C-3) and 3) the COVID-19 IgG/IgM Rapid Test Cassette (Whole Blood/Serum/Plasma) from Orient Gene / Healgen (Test lot 2003260), utilizing both the spike and the nucleocapsid protein. All three tests are based on immunochromatography for detection of IgG and IgM specific to SARS CoV-2 in human whole blood (venous and fingerstick) serum or plasma. We performed the tests following the manufacturers’ instructions. Each sample was tested by one test and readout (positive/negative) interpreted by two operators in parallel.

### PRNT 50

PRNT50 was used as a reference for this study, because neutralization assays are the standard for CoV serology. We tested serum and plasma samples for their neutralization capacity against SARS-CoV-2 (German isolate; GISAID ID EPI_ISL 406862; European Virus Archive Global # 026V-03883) by plaque-reduction neutralization test (PRNT50) as previously described (*Okba et al, EID, 2020*).

### Statistical Analysis

The outcome of commercial testing was correlated to functional antibody measurements to assess likelihood of predicting protective antibody responses. The results of the different ELISAs and RDTs were correlated to those detected by PRNT, as the gold standard for CoV serology at various time points after symptom onset. Results were analyzed using GraphPad Prism version 8 (https://www.graphpad.com) and sensitivity/specificity were calculated.

## Results and discussion

### 1. Population screening

The specificity of serological tools detecting antibodies against SARS CoV-2 might be hampered by the presence of antibodies against other circulating coronaviruses in the population, and thus testing for cross reactivity is crucial. In population screening during an early pandemic phase, a highly specific assay is required, to assure an acceptable positive predictive value in populations with a low sero-prevalence ^3^. This condition was met for all but one of the ELISA assays and the Liaison analyzer (Suppl table 1 and 2), when testing serum samples from persons exposed to a range of viruses. For the rapid tests, specificity varied more widely (Suppl table 3), using a restricted sample set.

The limited knowledge on antibody kinetics in emerging virus infections is always a challenge. A recent study ^3^ did show that in both hospitalized patients and patients with mild disease, seroconversion rates reach 100% after 10-14 days, and that antibody levels may correlate with clinical severity ^2,3^. This is in line with Middle East Respiratory Syndrome coronavirus (MERS-CoV) infection, in which antibody responses varied depending on disease severity, with mild and asymptomatic infections resulting in weaker immune responses ^4^. Therefore, for meaningful interpretation and extrapolation of population serology studies, sufficient follow-up samples from persons with mild and asymptomatic disease should be included. This patient group is underrepresented in the currently available validation data.

The golden standard for determining antibodies against coronaviruses up to present is virus neutralization, which detects antibodies capable of neutralizing SARS CoV-2 ^2^. Comparing the results of the available assays to our in-house plaque reduction neutralization assay (PRNT50) in Figure 1A-D shows a high sensitivity of the Wantai Ig assay using the RBD as coating antigen and detecting total immunoglobulins. For population-based serology, assays measuring long-lived IgG isotype antibodies or memory IgA antibodies are preferable, once these are expected to be present (from around 14 days post onset of disease). Both IgG and IgA assays in our comparison (Liaison and Euroimmun, Fig 1B,C) clearly demonstrate this, and especially the high-throughput Liaison system will be useful in population screening. However, both the testing in patients with subclinical disease and longer term follow-up of patients is needed to better understand the relationship between the antibody measurements and neutralizing antibodies. Multiplex platforms allowing differentiation between isotypes in a single serum sample will be an add-on to this question.

**Figure 1.**
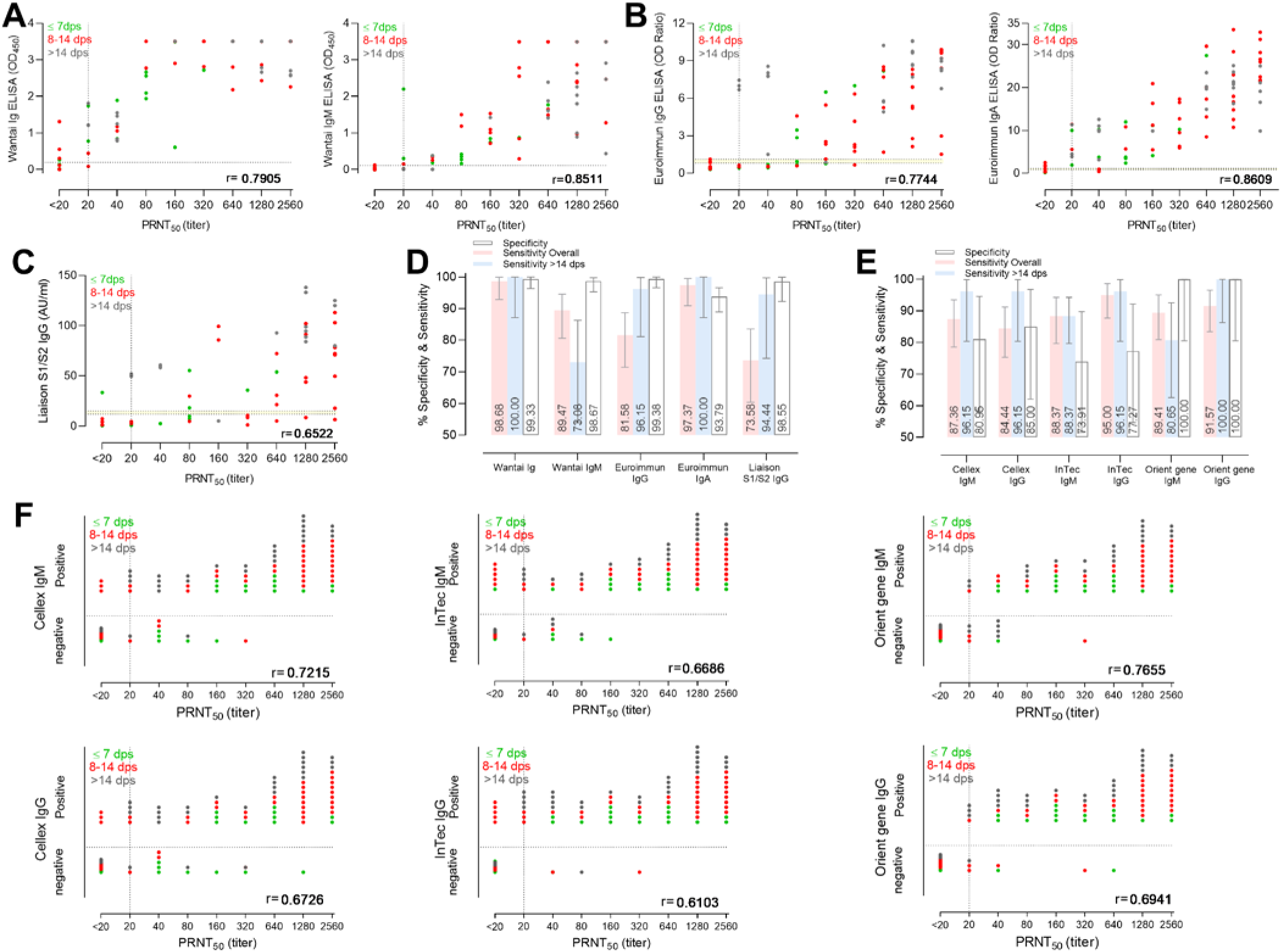
Performance of various commercial serological assays for detection of SARS-CoV-2-specific antibodies. Correlations of SARS-CoV-2 neutralizing antibody titers tested by a plaque reduction neutralization assay (PRNT50) to antibodies detected by A) Wantai Ig and IgM ELISAs, B) Euroimmun IgG and IgA ELISAs, C) DiaSorin Liaison XL S1/S2 IgG chemiluminescence immunoassay F) IgM and IgG Rapid diagnostic tests from Cellex, InTec and Orient gene. D,E) Specificities and sensitivities of various platforms tested. Dotted lines indicate the cut-off of each assay as indicated by the manufacturer: Wantai Ig ELISA, 0.19 OD; Wantai IgM ELISA, 0.105 OD, Euroimmun ELISAs, borderline 0.8–1.1 OD ratio (yellow shaded area), positive >1.1 OD ratio; Liaison S1/S1 IgG, borderline 12–15 AU/ml (yellow shaded area) and positive >15 AU/ml.

The observation that all samples testing positive in the Wantai Ig ELISA with a signal > 2 had high levels of neutralizing antibodies suggests that - using a cut-off-this assay could be used to indicate presence of protective antibodies, although a true correlate of protection remains to be determined. For further quantification, linearity of the ELISA results compared to the neutralizing antibody titers was assessed. Here, the Euroimmun IgA assay shows a good quantitative relationship, specifically once neutralizing titers were higher than 80 (PRNT50 units), upon which the Wantai Ig assay becomes non-linear. As IgA testing will detect both early and memory IgA responses, it will be a useful addition. The possible relevance of quantitative antibody measurements will need to be assessed when results of longer term patient follow up studies become available.

In addition to specialized ELISA assays used in laboratory settings, a wide range of rapid tests has been put on the market, triggering the question whether they can be used for widespread testing. The selected rapid tests in our evaluation provide qualitative (yes/no) results which does not allow quantification or the definition of a cut-off for neutralization (Figure 1E,F). Although the Orient gene rapid test, showing high specificity (Suppl. Table 3), seems useful for population screening, additional validation in patients with mild disease is required to assess the possible under-ascertainments. An additional drawback of rapid testing outside controlled laboratory settings is registration the poor of the results and the possible risk that people will interpret a positive test outcome as a measure of protection.

### 2. Patient diagnostics

Serological testing to support clinical diagnostic work-up is mostly requested in hospitalized patients for example when SARS CoV-2 RNA diagnostic testing remains negative in a patient despite a strong clinical suspicion, for patients whose samples during the acute phase were not collected, or in patients with low viral loads awaiting decision to end isolation measures. In these patients, the onset of disease is known, as well as the clinical phase of disease. Usually these serological results are interpreted by laboratory staffs and there is a possibility to test follow-up sera or perform confirmation serological testing. When carefully taking into account the performance characteristics, the validated commercial ELISAs can be applied in patient care, but the Liaison platform will lack sensitivity at early time-points. For all assays tested, it is clear that the use of rapid tests during triage - as has been suggested in some marketing campaigns - should be discouraged as the sensitivity of serology in early stage infection is too low.

In conclusion, based on our assessment to date, the RBD ELISA had optimal characteristics for use in both population-level serological testing and patient diagnostics, including the potential to set a cut-off indicating high levels of protective antibodies. Although some rapid tests could be used for population level antibody screening, their performance is not robust enough for over the counter personalized testing.

## Data Availability

all data is included in the manuscript

## Acknowledgements

Zain Abdel-Karem Abou-Nouar, Sandra Scherbeijn are acknowledged for excellent technical assistance.

## Supplementary tables

**Table 1.** Sample panel used to validate the performance of the ELISA/Liaison and the antibody RDT assays for SARS-CoV-2

| Sensitivity |  |  |  |  |
| --- | --- | --- | --- | --- |
| Country | Sample source | Infection | No. samples Liaison/ELISA (RDT*) | Post symptom onset range |
| Netherlands | RT-PCR confirmed SARS-CoV-2 | Mild/Moderate | 0/15 (15) | 1-24 |
|  |  | Severe | 55/78 (78) | 1-43 |
| Specificity |  |  |  |  |
| Netherlands | Non-CoV respiratory infections* |  |  |  |
|  |  | Adeno virus | 0/5 (1) | 2-4weeks |
|  |  | HMPV | 3/9 (3) | 2-4weeks |
|  |  | Flu A | 8/10 (4) | 2-4weeks |
|  |  | Flu B | 5/6 (4) | 2-4weeks |
|  |  | RSV A | 5/5 (4) | 2-4weeks |
|  |  | RSV B | 4/4 (4) | 2-4weeks |
|  |  | CMV | 1/4 (2) | 2-4weeks |
|  |  | EBV | 1/7 (3) | 2-4weeks |
|  |  | *M.pneumoniae* | 1/1 (1) | 2-4weeks |
|  |  | Rhinovirus | 9/9 (2) | 2-4weeks |
|  |  | Bocavirus | 0/2 (0) | 2-4weeks |
|  |  | Parainfluenza 1/3 | 0/4 (0) | 2-4weeks |
|  |  | Enterovirus | 0/2 (0) | 2-4weeks |
| Netherlands | hCoV infections |  | 9/19 (6) | 2-4weeks |
|  |  | HCoV 229E |  |  |
|  |  | HCoV-NL63 | 8/18 (7) | 2-4weeks |
|  |  | HCoV-OC43 | 21/39 (9) | 2-4weeks |
| Netherlands |  | MERS-CoV | 1/3 (3) | 2-4weeks |

\* numbers were limited due to RDT kit availability

**Table 2.**
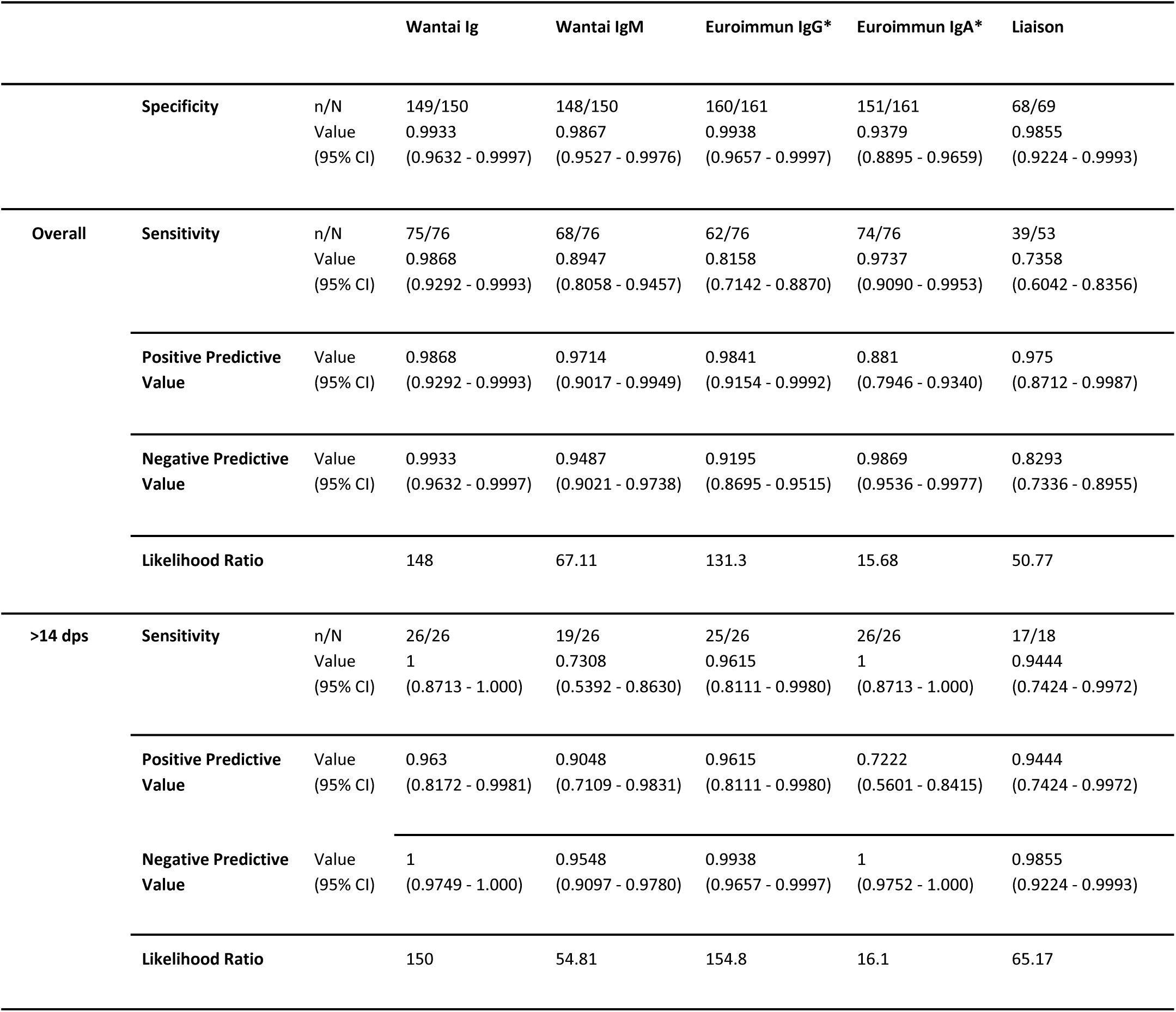
A summary of the performance characteristics of the Wantai Ig and IgM ELISA, Euroimmun IgG and IgA ELISA and DiaSorin Liaison platform.

The outcome of the commercial assays was compared to in-house PRNT50. For sensitivity calculations therefor only the PRNT50 positive samples were used for the calculations.

**Table 3.** Sensitivity and specificity of the three tested RDTs (CI 95%) compared to PRNT_50_ positivity: whole sample set and samples >14 days post symptom onset (borderline PRNT50 values counted as positive).

|  | Cellex (n=93) |  | InTec (n=93) |  | Orient gene (n=90) |  |
| --- | --- | --- | --- | --- | --- | --- |
|  | IgM | IgG | IgM | IgG | IgM | IgG |
| **Compared to PRNT<sub>50</sub> positivity** |  |  |  |  |  |  |
| **Sensitivity-overall** | **87.36%**<br>(78.50% to 93.52%) | **84.44%**<br>(75.28% to 91.23%) | **88.37%**<br>(79.65% to 94.28%) | **95.00%**<br>(87.69% to 98.62%) | **89.41%**<br>(80.85% to 95.04%) | **91.57%**<br>(83.39% to 96.54%) |
| **Sensitivity >14 DPO**<br>**(n=25)** | **96.15%** (80.36% to 99.90%) | **96.15%** (80.36% to 99.90%) | **88.37%** (79.65% to 94.28%) | **96.15%** (80.36% to 99.90%) | **80.65%** (62.53% to 92.55%) | **100.00%** (86.28% to 100.00%) |
|  | **IgM and IgG**<br>96.15%<br>(80.36% to 99.90%) |  | **IgM and IgG**<br>96.15%<br>(80.36% to 99.90%) |  | **IgM and IgG**<br>100.00%<br>(86.28% to 100.00%) |  |
| **Specificity -overall\*** | **80.95%**<br>(58.09% to 94.55%) | **85.00%** (62.11% to 96.79%) | **73.91%** (51.59% to 89.77%) | **77.27%** (54.63% to 92.18%) | **100.00%** (80.49% to 100.00%) | **100.00%** (80.49% to 100.00%) |

\* number of sera tested for specificity based on availability of test kit; n=44, 64 and 9 respective of Cellex, inTec and Orient Gene

The outcome of the RDTs was compared to in-house PRNT50. For sensitivity calculations therefor only the PRNT50 positive samples were used for the calculations.

